# Machine Learning-Based Identification of Sickle Cell Disease Subphenotypes in Clinical Trial Data

**DOI:** 10.1101/2025.06.01.25328537

**Authors:** Wei Xiao, Patricia Oneal, Menglun Wang, Nihar J. Mehta, Qi Liu, Rongmei Zhang, Susan Perrine, Qin Ryan

## Abstract

Sickle Cell Disease (SCD) is a rare autosomal recessive disorder caused by a point mutation producing abnormal hemoglobin S, leading to deformed red blood cells and a wide range of clinical manifestations, including pain crises, organ damage, and an increased risk of infection. These devastating complications often result in significant morbidity and early mortality, presenting significant therapeutic challenges. Currently, there is a lack of clinically validated predictive tools to assess individual SCD patients’ prognoses and therapeutic responses. This is largely due to the complexity and variability of the clinical manifestations, which vary widely among patients. As a result, there remains an unmet need for a systematic approach to SCD disease subphenotype classification that can guide and tailor therapeutic strategies, predict outcomes, and improve patients’ lives. Over a decade ago, two clinical subphenotypes in SCD were proposed based on literature and clinical observations. However, this concept has not been applied or explored in the design of clinical trials (CT). Recent advances in machine learning (ML) applications in medicine, and growing availability of SCD clinical trial data evaluating therapeutics which target different pathophysiologic aspects of the disease, provides opportunity to enhance understanding of therapeutic responses within SCD populations. Applying ML techniques to a large CT database could support development of robust disease models capable of identifying and validating disease subphenotypes, with potential to predict outcomes to specific therapies based on mechanism of action and to optimize care in SCD.

In this study, we constructed a comprehensive database comprising 3,551 patients with SCD from 16 clinical trials that supported therapeutic approvals for SCD. Using this database, we applied a machine learning pipeline to develop a rule-based classification method, which identified two distinct clinical subphenotypes of SCD: the Vaso-occlusive Primary (VP) subphenotype, primarily characterized by a higher frequency of vaso-occlusive pain crises, and the Hemolytic Dominant (HD) subphenotype, characterized by chronic hemolysis and its associated complications. Biomarker comparisons demonstrated that the VP subphenotype was associated with a significantly higher annual rate of vasoocclusive crisis events, significantly higher levels of total and fetal hemoglobin, and leukocytosis, while the HD subphenotype exhibited significantly higher levels of hemolysis-related biomarkers of indirect bilirubin. The biomarker profiles were validated using an independent clinical trial dataset, which confirmed these two subphenotypes in SCD.

Our study demonstrated that the integration of ML with disease pathophysiology enables robust identification of clinically meaningful subphenotypes of SCD from an international clinical trial database. This approach provides a basis for developing predictive disease models, which may optimize treatment strategies and improve patients’ outcomes. Further, our methodological framework offers a scalable model for application to identify subsets in other rare genetic diseases.

## Introduction

Sickle Cell Disease (SCD), a group of autosomal recessive disorders caused by mutations in the beta globin gene of adult hemoglobin (HbA, α_2_β_2_), includes sickle cell anemia (HbSS), HbSC, HbS-β-thalassemia, and other HbS-related hemoglobinopathies (1–5). Due to the genetic and pathophysiological variability across these subtypes, clinical manifestations and associated laboratory biomarkers vary significantly from patient to patient (6–15). This heterogeneity creates an unmet need for predictive phenotypic disease models to optimize potential treatments, estimate short- and long-term disease outcomes, and guide therapeutic development for subjects with SCD to those more likely to benefit from therapies which address their phenotype.

Despite decades of research, individuals with SCD continue to experience poor health outcomes and lower life expectancy than the unaffected population (26, 27). Supportive treatments have been developed for identified risks such as prophylactic antibiotics, targeted vaccines, red blood cell transfusions, pain management, hydroxyurea therapy, and, for those with donors, stem cell transplantation (16–23). Although a few therapeutics were recently approved, clinical trials of well-designed therapies have faced significant challenges related to the variability of disease complications (24, 25).

SCD is characterized by chronic hemolytic anemia and recurrent vaso-occlusive crises, leading to a wide range of complications. However, prognostication remains difficult due to gaps in our current understanding of disease progression and the lack of robust, individualized predictive models (26–28). Although a number of laboratory and genetic modifiers have been identified (67–81), biological and clinical prognostic factors that reliably predict outcomes or survival in SCD have not been studied in large clinical trial datasets.

Previous reports have hypothesized two major SCD subphenotypes based on predominance of two pathophysiologic mechanisms-cell adhesion to the vasculature causing vaso-occlusive crises or hemolysis and endothelial dysfunction, respectively (29–31). However, distinguishing between these subphenotypes in practice is complicated by overlapping clinical presentations, and this hypothesis has not been evaluated in clinical trials.

In this study, we applied machine learning (ML) modeling and computational analyses to a large, curated clinical trial database of patients with SCD, to identify distinct disease subphenotypes. This approach aims to enhance understanding of disease heterogeneity, evaluate how specific subphenotypes respond to therapies directed to a predominant pathophysiology, and provide a foundation for predictive algorithms that can inform future clinical trial design. Moreover, our ML pipeline offers a scalable framework that may also benefit research in other rare genetic diseases.

## Methods

### Database

The clinical trial baseline database utilized in this study was constructed from 16 clinical trials in SCD submitted to the U.S. Food and Drug Administration (FDA) between 2017 to 2021. Data were obtained from 3,551 patients with baseline clinical data encompassing demographics, genotypes, disease characteristics, medical history of SCD-related clinical complications, and laboratory test results collected prior to any trial interventions (Figure 1). Most baseline data were obtained within 12 months before enrollment. Notably, enrolled patients were permitted to be on a stable dose of hydroxyurea before enrollment in these trials. Additionally, data from an independent study, the Multicenter Study of Hydroxyurea in Sickle Cell Anemia (MSH) (32), provided by National Institutes of Health, National Heart, Lung, and Blood Institute, which randomized participants to hydroxyurea or placebo, were used for model evaluation.

**Figure 1.**
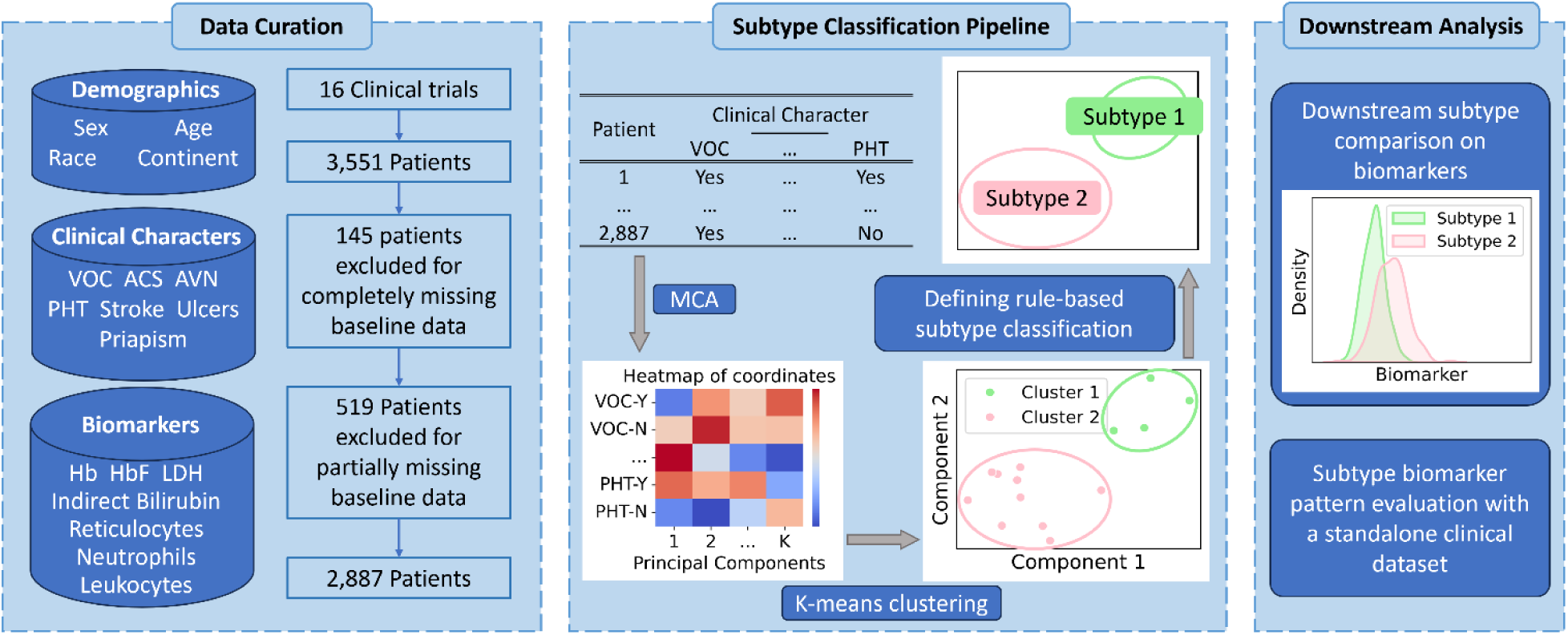
Data processing workflow. 1. Data Curation: SCD patients’ demographics, clinical characteristics, and biomarkers were extracted from multiple clinical trials submitted to the FDA. Patients with insufficient data were excluded from the analysis. 2. Subtype Classification Pipeline: Multiple Correspondence Analysis (MCA) was applied to the clinical features, obtaining coordinates for each category of clinical features across the principal components. K-means clustering was then performed on these coordinates, resulting in the classification of clinical features into two distinct clusters. 3. Downstream Analysis: Based on the clustering results, two subphenotypes of SCD were defined, and the differences in biomarkers between the two subphenotypes were subsequently compared.

### Variable Selection

For disease subphenotype classification, we selected seven clinical features commonly associated as SCD complications: vaso-occlusive pain crisis (VOC), acute chest syndrome (ACS), avascular necrosis (AVN), pulmonary hypertension (PHT), stroke, leg ulcers (ulcers), and priapism. Each feature was recorded as a binary indicator at baseline for each patient: “Yes” if the feature had occurred at least once and “No” if it had never occurred during the entire baseline data collection period. Patients with data missing on any of these variables were excluded from the analysis, resulting in a final dataset of 2,887 patients, representing 81.3% of the total database.

### Clustering Analysis

To identify potential disease subphenotypes, we conducted Multiple Correspondence Analysis (MCA) on a dataset comprising 2,887 patients with complete data for seven binary clinical features: VOC, ACS, AVN, PHT, stroke, ulcers, and priapism. MCA (33) is a dimensionality reduction technique suitable for categorical data, transforming the original variables into a set of principal components that capture the underlying structure of the data. In our analysis, each distinct category (“Yes” or “No”) of the clinical features was represented in the transformed seven-dimensional space as seven principal components, resulting in seven coordinates corresponding to 14 features. Subsequently, we applied K-means clustering (34) to the coordinates derived from the first five principal components, specifying two clusters as the target (Figure 1).

### Downstream Primary and Secondary Analysis

We defined two SCD subphenotypes based on clustering results and assigned patients accordingly. In the primary analysis, 2,887 patients from our clinical trial database were categorized into these subphenotypes. To assess baseline differences, we selected several disease-relevant laboratory biomarkers, including hemoglobin (Hb), fetal hemoglobin (HbF), lactate dehydrogenase (LDH), reticulocytes, indirect bilirubin, neutrophils, and leukocytes (Figure 1). Additionally, the annual rate of VOC was included as a numerical variable. The Mann-Whitney U test (35) was employed for continuous variables, while the chi-square test (36) was used for the discrete variable of annual VOC rate. For secondary analysis, we applied our rule-based subphenotype classification to individual patient data (n=299) from an independent SCD clinical trial, the Multicenter Study of Hydroxyurea in Sickle Cell Anemia (32). Similar statistical methods were utilized to evaluate biomarkers and the annual VOC rate in this cohort to validate the primary analysis.

## Results

### Clinical Feature Clustering

Previous research has hypothesized that sickle cell disease comprises two primary subphenotypes driven by distinct underlying mechanisms (28–29). One subphenotype, associated with hemolysis and low steady-state hemoglobin levels, is more likely to experience complications related to nitric oxide consumption by free hemoglobin, such as pulmonary hypertension (PHT), stroke, leg ulcers (ulcers), and priapism. The other subphenotype is characterized by vaso-occlusive events, including pain crisis (VOC), acute chest syndrome (ACS), and avascular necrosis (AVN), and is associated with higher steady-state leukocyte counts and relatively higher hemoglobin levels, and associated with increased red cell adhesion to endothelium (30).

To investigate this hypothesis in clinical trial data, we curated a database comprising 3,551 patients from 16 clinical trials that supported drug approvals for treatment of SCD. After excluding 664 patients with incomplete clinical feature data, 2,887 patients (81.3% of the database) remained for analysis.

Demographic characteristics of these patients are summarized in Supplementary Table 1. The clinical features were selected to assess potential stratification within this trial dataset that align with the hypothesized subphenotypes (30), potentially elucidating variations in disease severity and complications. The data processing workflow is illustrated in Figure 1.

Disease variable assessment was conducted based on clinical relevance and data availability, resulting in the selection of seven primary clinical features as subjects for our analytical pipeline: VOC, ACS, AVN, PHT, stroke, ulcers, and priapism. Although jaundice is a common clinical feature of SCD, it was excluded due to significant missing data.

After several methodology explorations, to analyze binary clinical features (“Yes” or “No”), we applied Multiple Correspondence Analysis (MCA) (33), a dimensional reduction technique for categorical data. The MCA provided percentage contributions of each clinical feature category to the principal components (Supplementary Figure 1). We combined the “Yes” and “No” categories for each clinical feature by percentage contributions to clarify their significance within each principal component (Figure 2A). The relatively even weight of each feature in explaining the data variance (Figure 2B) led to a rearrangement of principal components, revealing two distinct modules (Figure 2A). Notably, PHT, stroke, ulcers, and priapism clustered together, suggesting a common underlying mechanism. Interestingly, these clinical features were related to the hemolytic subphenotype in previous reports by Kato et al (28–29, 37).

**Figure 2.**
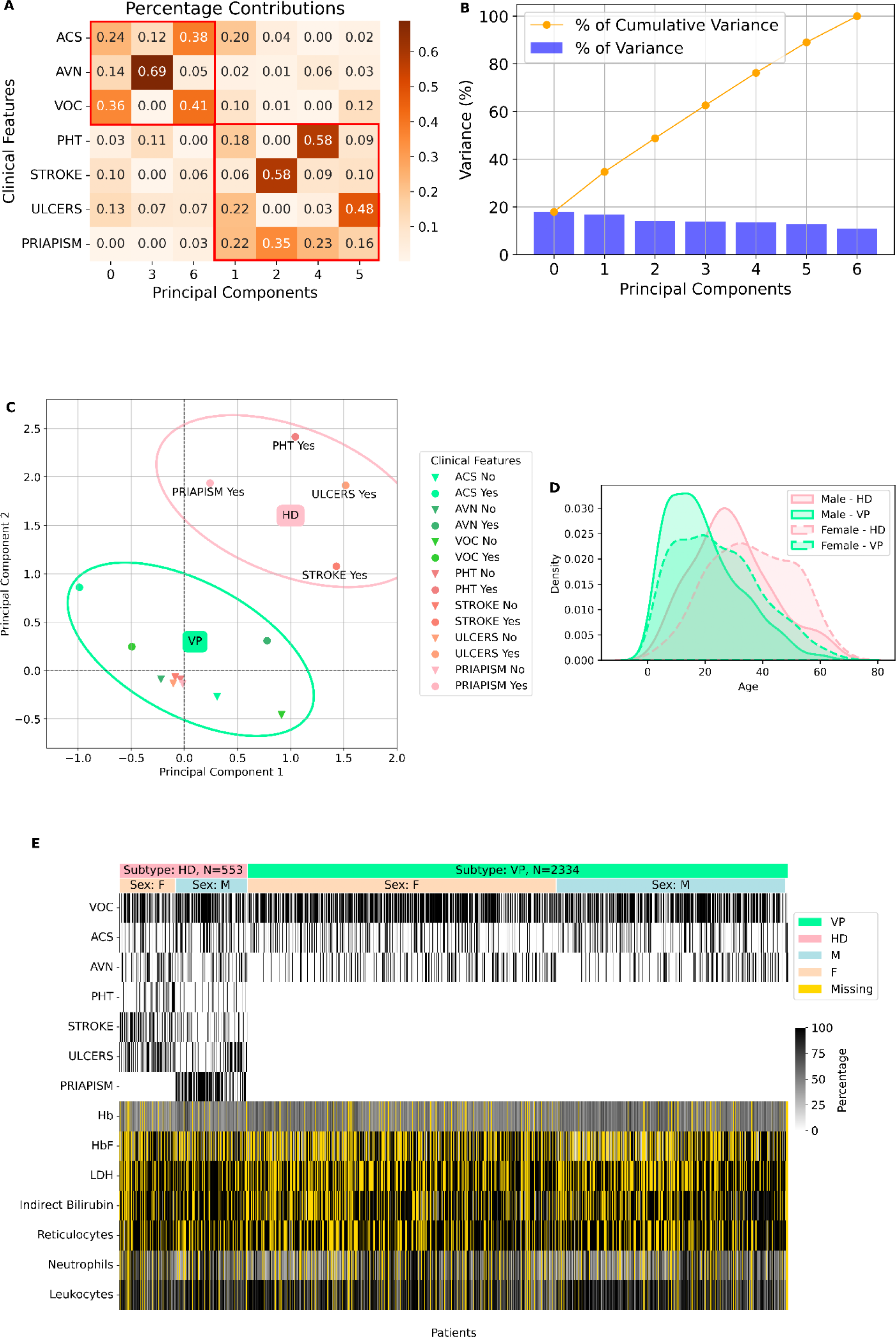
Identification of the two subphenotypes of SCD based on clinical features. (A) The percentage contributions of each clinical features to each principal component (the two categories, “Yes” and “No”, of each clinical feature are combined for clearer pattern visualization). (B) The proportion of data variance explained by each principal component. (C) The projection of both categories of all clinical features along the first two principal components, with circles highlighting the two subphenotypes identified by K-means clustering. The green circle represents the Vaso-Occlusive Primary (VP) subphenotype, while the pink circle represents the Hemolysis Dominant (HD) subphenotype. (D) Age distributions across combined subsets of gender and subphenotype. (E) A heatmap visualizing clinical features and laboratory biomarkers over the baseline dataset patients with SCD. The first two rows represent the SCD subphenotype and sex of each patient, with patient ordering prioritized by SCD subphenotype, followed by sex, and then by age. Clinical and laboratory features, shown in subsequent rows, are normalized to a 0-100 scale to standardize across diverse measurement ranges (reference to laboratory upper normal limit of healthy people, Supplementary table 5). The yellow color in the heatmap represents patients with missing laboratory biomarkers.

Subsequently, we utilized the coordinates of the first five principal components, which accounted for 76.25% of the variance, and performed K-means clustering specifying two clusters. The clustering results (Figure 2C) indicated that the “Yes” categories of PHT, stroke, ulcers, and priapism grouped into one cluster, while the “No” categories of these features, along with VOC, ACS and AVN, formed a second cluster. These findings support the existence of two SCD subphenotypes: one associated with hemolysis-related clinical features, and the other without.

### Subphenotype Define and Comparison

Based on the K-means clustering results of seven binary clinical features, we identified two SCD subphenotypes. The Vasoocclusive Primary (VP) subphenotype was characterized by the absence of PHT, stroke, ulcers, or priapism. In contrast, the Hemolysis-Dominant (HD) subphenotype was defined by the presence of at least one of these features. Among the 2,887 patients analyzed in our rule-based subphenotype classification, 2,329 were classified as VP and 552 as HD (Figure 2E).

The distribution of clinical features and laboratory biomarkers analyzed for ML subtyping at the subject level is depicted in Figure 2E. In this figure, subjects were organized by HD and VP subphenotypes, followed by sex (female and male), and then by ascending age. The age ranges were as follows: HD females and HD males, 3-67 years; VP females, 1-70 years; and VP males, 0-65 years. A difference of sex and age distributions between the two subphenotypes was found. Females were significantly older than males in both subphenotypes and overall, patients in the HD subphenotype were generally older than those in the VP subphenotype (Figure 2D and Supplementary Figure 2)

To elucidate the definitions of the two subphenotypes, seven clinical features used for classification were visualized in Figure 2E. While PHT, stroke, ulcer and priapism were exclusively presented for the HD subphenotype, VOC, AVN, and ACS were populated in both subphenotypes with various prevalence (Table 2E). Additionally, laboratory biomarkers were displayed in Figure 2E, with all values normalized to a 0-100 scale. Given the strong association between age and biomarker values, age-dependent scaling for each biomarker was applied during the normalization to address associations between age and biomarker levels (Supplementary Table 3).

We compared numerical differences in laboratory biomarkers and a numerical clinical feature, the annual rate of VOC, between the two subphenotypes. As depicted in Figure 2E and Supplementary Table 4, a notable proportion of subjects had missing data for certain laboratory biomarkers. To address this, we conducted separate analyses for each biomarker, excluding only those patients who lacked data for the specific biomarker under consideration. The number of patients involved in each analysis is listed in Figure 3.

**Figure 3.**
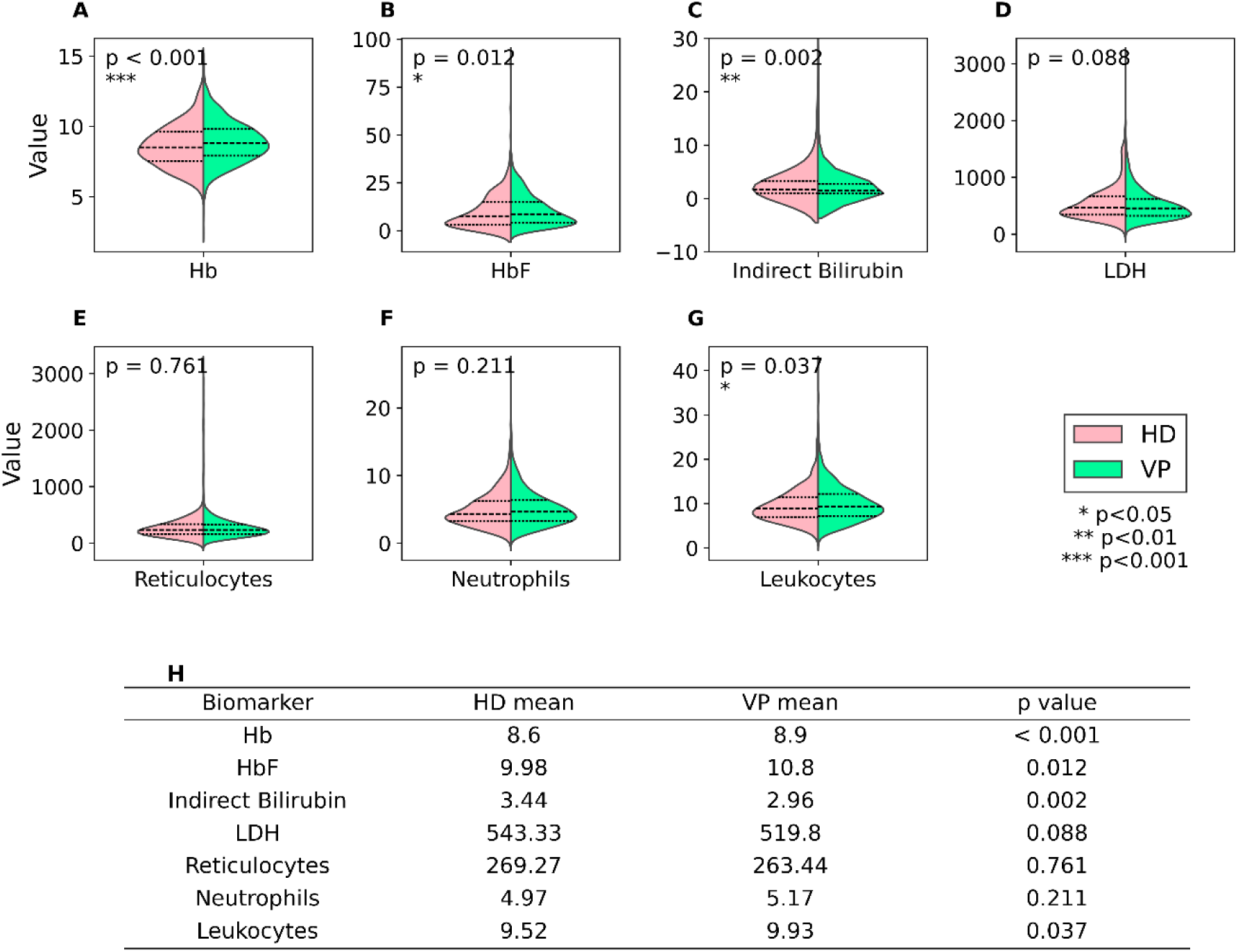
Violin plots showing differences in laboratory biomarkers between the VP and HD subphenotypes at baseline of the Clinical Trial population. In each plot, the median value is represented by a dashed line, and the 75th and 25th percentiles are indicated by doted lines. (A) Hemoglobin levels (g/dL, N_total_=2,583, N_VP_=2,084, N_HD_=502). (B) Fetal hemoglobin levels (%, N_total_=1,671, N_VP_=1,330, N_HD_=341). (C) Indirect bilirubin (mg/dL, value has been zoomed in for better visualization. N_total_=1,941, N_VP_=1,541, N_HD_=400). (D) Lactate dehydrogenase (LDH) levels (U/L, N_total_=1,623, N_VP_=1,295, N_HD_=328). (E) Reticulocyte counts (10^9^/L, N_total_=1,914, N_VP_=1,528, N_HD_=386). (F) Neutrophils (10^9^/L, N_total_=2,365, N_VP_=1,901, N_HD_=464). (G) Leukocytes (10^9^/L, N_total_=2,545, N_VP_=2,050, N_HD_=495). (H) A table showing mean biomarker levels in VP and HD subphenotypes, with p-values from the Mann-Whitney U test indicating statistical differences between the two groups. VP: Vasoocclusive Primary subphenotype; HD: Hemolysis-Dominant subphenotype.

First, we compared baseline hemoglobin (Hb) levels between the VP and HD subphenotypes. VP patients exhibited a mean total Hb of 8.9 g/dL compared to 8.6 g/dL in HD patients (Mann-Whitney U test, p < 0.001, Figure 3A, 3H). Similarly, fetal hemoglobin (HbF) values were slightly, but significantly, higher in the VP group, with a mean of 10.8%, compared to 9.98% in the HD group (Mann-Whitney U test, p = 0.012, Figure 3B, 3H). Second, small, but significantly, lower baseline levels of indirect bilirubin were found in VP patients compared to HD patients, with mean values of a 2.96 mg/dL and 3.44 mg/dL, respectively (Mann-Whitney U test, p = 0.002, Figure 3C, 3H). As elevated indirect bilirubin is a known marker of hemolysis, its higher levels in the HD subphenotype supports the rule-based classification. The ranges of each laboratory values are shown in Figure 3. Despite overall higher levels of LDH exhibited in the database, mean differences between the two subphenotypes were not statistically significant (p = 0.089), nor were mean differences in absolute reticulocytes (p = 0.76) or absolute neutrophils (p = 0.21) (Means are tabulated in Figure 3H. Mann-Whitney U test, Figure 3D-F, and 3H). However, LDH levels in the HD subphenotype exhibited a bimodal distribution, with a subset of HD patients demonstrating substantially higher LDH levels (Figure 3D, detail in Supplementary Figure 3). This distribution indicates heterogeneity within the HD subphenotype. Leukocyte counts were significantly higher in the VP subphenotype (mean: 9.93 x 10^9^/L) than in the HD patients (mean: 9.52 x 10^9^/L, p = 0.037, Figure 3G, 3H). The higher leukocyte count in VP patients may be related to inflammatory responses associated with vaso-occlusive crises following vascular obstruction and tissue ischemia (37, 38).

Our baseline CT data demonstrate a distinct difference in the annual rate of VOC events between the two subphenotypes. Patients with the VP subphenotype experienced a significantly higher rate of painful crises compared to those with the HD subphenotype (chi-square test, p = 0.012). As shown in Figure 4, a markedly higher number of subjects classified in VP subphenotype experienced annual numbers of vaso-occlusive crises from 1-5/year, despite sizeable proportions of patients in both groups (VP: 39.5%, HD: 50.0%) who did not report any painful crises during the baseline data collection periods during the trials. Additionally, 1,072 patients (37.1% of the cohort) were excluded from this analysis due to missing data on baseline annual rate of VOC events. Consequently, the proportion of patients with at least one VOC event, as determined by this variable, differs from the binary clinical feature “VOC” used to define the VP subphenotype (Supplementary Table 2). In summary, the VP subphenotype is characterized by a higher annual rate of VOC events, increased total hemoglobin levels, and higher steady-state leukocyte counts, whereas the HD subphenotype is marked by elevated hemolysis-related biomarkers, such as indirect bilirubin and LDH (Table 1).

**Figure 4.**
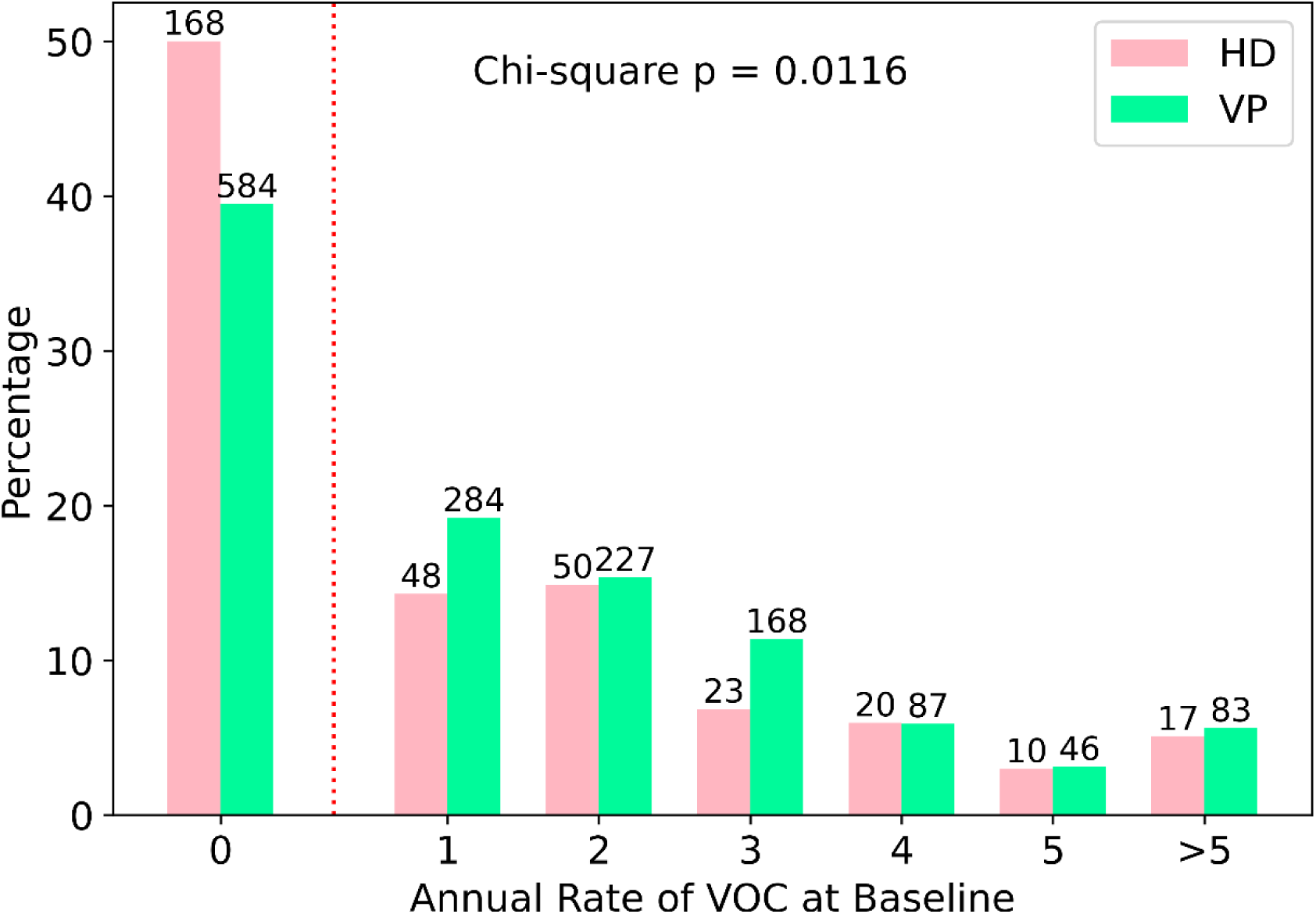
The figure shows the percentage of patients with various annual rates of VOC events at baseline, stratified by subphenotype. The patient counts in each group are labeled at the top of the bar. N_total_=1,815, N_VP_=1,479, N_HD_=336. The red dotted line divided patients without VOC events and patients with VOC events. VP: Vasoocclusive Primary subphenotype; HD: Hemolysis-Dominant subphenotype.

**Table 1.** Summary of subphenotyping analyses. Comparison of laboratory biomarkers and the VOC annual rates between VP and HD subphenotypes in the SCD clinical trial database and the data of MSH study. The table shows the trend of differences for each feature, with statistical significance indicated by asterisks (*). VP: Vasoocclusive Primary subphenotype; HD: Hemolysis-Dominant subphenotype.

|  | Primary Analysis<br>SCD Clinical Trial Database |  | Secondary Analysis<br>Data of MSH Study |  |
| --- | --- | --- | --- | --- |
|  | VP | HD | VP | HD |
| Hb | Higher* | Lower* | Higher* | Lower* |
| HbF | Higher* | Lower* | Higher* | Lower* |
| Indirect Bilirubin | Lower* | Higher* | Lower | Higher |
| LDH | Lower | Higher | - | - |
| Reticulocytes | Same | Same | Same | Same |
| Neutrophils | Same | Same | Same | Same |
| Leukocytes | Higher* | Lower* | Same | Same |
| Annual rate of VOC | More* | Less* | Same | Same |

### Evaluation of Subphenotype Definition on the Multicenter Study of Hydroxyurea in Sickle Cell Anemia

To verify our subphenotype definition, we conducted a comparable analysis using data from an independent trial, Multicenter Study of Hydroxyurea (MSH). The MSH study was a 1:1 randomized, double-blind, placebo-controlled clinical trial conducted from 1992-1995, enrolling 299 subjects with sickle cell anemia and aged 18 and older. The study evaluated the efficacy and safety of hydroxyurea, leading to its approval for treating adults with SCD in March 1998.

At baseline, all MSH participants were treatment -naïve, having received only supportive care such as RBC transfusion and pain management. Therefore, the recorded baseline clinical features and laboratory biomarkers should reflect natural history of SCD in the absence of disease modifying therapy. The MSH trial collected data on five clinical features: ACS, AVN, stroke, ulcers, and priapism. PHT data were not recorded, possibly due to the limited access for definitive diagnostic tools like right heart catheterization at the time. Additionally, all (100%) participants had baseline VOC events recorded, as eligible patients were required to have had at least three VOCs during the year preceding enrollment. Demographic characteristics of the MSH cohort are summarized in Supplementary Table 5.

Initially, we attempted to input the five available clinical features from 299 patients in the MSH study using MCA and applied K-means clustering on coordinates of the first few principal components. However, clustering based on the first three to five principal components only grouped one or two of the three clinical features—stroke, ulcers, and priapism—together. This limited clustering is likely attributable to the small sample size and two unimputable clinical features, PHT (100% absence), and VOC (100% presence) in the MSH dataset. Consequently, we methodically determined that subphenotype definitions derived from our previous ML-modeling, which utilized seven clinical features, could be suitable for assessing MSH study data.

Subsequently, we applied the previously established subphenotype definition to the MSH dataset using the five available clinical features. Patients were classified into two groups: the VP subphenotype, characterized by the absence of stroke, ulcers, or priapism at baseline, and the HD subphenotype, defined by the presence of at least one of these features. With this rule-based classification method, the 299 MSH patients were categorized into two groups: 166 VP patients and 133 HD patients. We then compared the same biomarkers and annual rate of VOC between these two subphenotypes.

Our analysis revealed that VP patients exhibited significantly higher Hb levels (mean: 8.69 g/dL) compared to HD patients (mean: 8.23 g/dL, Mann-Whitney U test, p = 0.0023, Figure 5A, 5G). Similarly, HbF showed the same trend as Hb, elevated in VP with mean HbF 5.75% compared to HD with mean HbF 4.25% (Mann-Whitney U test, p < 0.001, Figure 5B, 5G).

**Figure 5.**
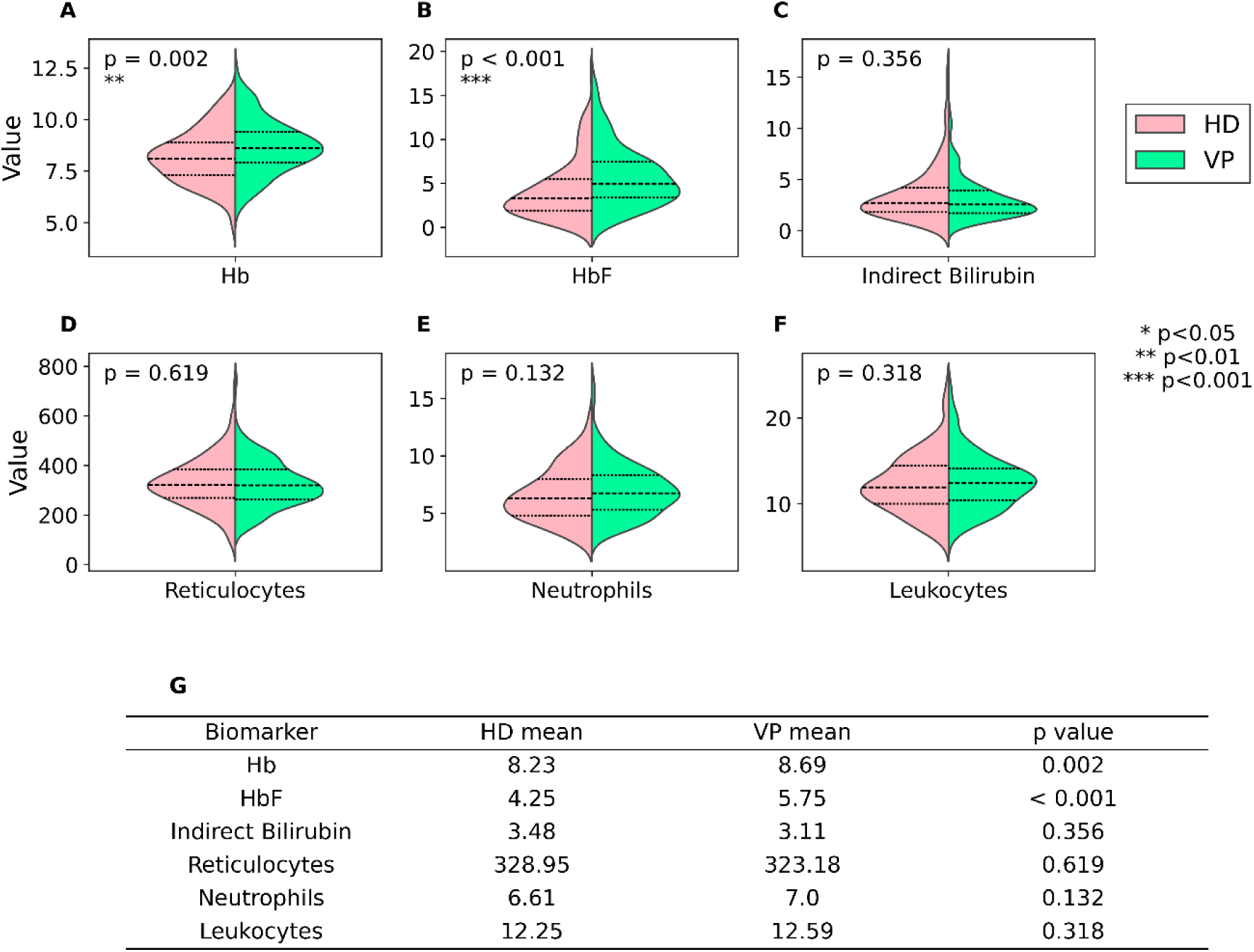
Violin plots showing differences in laboratory biomarkers between the VP and HD subphenotypes at baseline of MSH Study (N_total_=299, N_VP_=166, N_HD_=133). In each plot, the median value is represented by a dashed line, and the 75th and 25th percentiles are indicated by doted lines. (A) Hemoglobin levels (g/dL). (B) Fetal hemoglobin levels (%). (C) Indirect bilirubin (mg/dL). (D) Reticulocyte counts (10^9^/L). (E) Neutrophils (10^9^/L). (F) Leukocytes (10^9^/L). (G) A table showing mean biomarker levels in VP and HD subphenotypes, with p-values from the Mann-Whitney U test indicating statistical differences between the two groups. VP: Vasoocclusive Primary subphenotype; HD: Hemolysis-Dominant subphenotype.

In contrast, no statistically significant differences were found between the two subphenotypes in either indirect bilirubin (Mann-Whitney U test, p = 0.36, Figure 5C, 5G) or reticulocyte counts (Mann-Whitney U test, p = 0.62, Figure 5D, 5G). However, a trend toward higher indirect bilirubin levels in the HD subphenotype was noted. This lack of statistical significance may be attributed to the limited sample size of the MSH study and potential misclassification due to incomplete clinical feature data. Notably, LDH was not available in the MSH dataset.

Further comparison showed no significant difference in neutrophil counts (Mann-Whitney U test, p = 0.13, Figure 5E, 5G) and leukocyte counts (Mann-Whitney U test, p = 0.32, Figure 5F, 5G) between the two subphenotypes. Nonetheless, leukocyte counts tend to be higher in the VP subphenotype compared to the HD subphenotype (Figure 5F). Additionally, we assessed the annual rate of VOC at baseline between the subphenotypes. No significant difference was found (chi-square test, P = 0.92, Supplementary Figure 4).

### Summary of Biomarker and the Annual Rate of VOC in VP and HD Subphenotypes

Based on our downstream analyses in both the clinical trial database and the MSH study data, comparative patterns between the VP and HD subphenotypes for biomarkers and the annual rate of VOC, are summarized as presented in Table 1. In the primary analyses with the clinical trial database, the VP subphenotype exhibited significantly higher Hb, HbF, leukocytes and annual VOC rates. Conversely, the HD subphenotype showed significantly elevated indirect bilirubin and a trend toward higher LDH. These findings strongly supported our rule-based subphenotype classification derived from clinical feature clustering analysis.

In our secondary analyses using MSH data, the results from application of our rule-based subphenotype classification were generally consistent with our primary findings from the SCD clinical trial database (Table 1). Even in instances where differences did not reach statistical significance, the observed trends aligned with those identified in the primary analyses.

It is important to note that the baseline data from the MSH study represented the natural history of SCD, as participants were naïve to hydroxyurea and other disease modifying drugs at the time of enrollment. This provided a unique opportunity to validate our primary findings, which were derived from a mixed SCD patient population, some of whom had been exposed to hydroxyurea.

However, the MSH dataset has limitations, including a smaller sample size and certain unfeasible clinical feature data, which may affect the precision of subphenotype classification. Despite these constraints, it remains the most suitable available dataset for evaluating our subphenotype classification. We will discuss these limitations further in the discussion.

In conclusion, the clustering of clinical features, along with distinct patterns in biomarker levels and the annual rate of VOCs, supports the differentiation between vasoocclusive primary (VP) and hemolytic dominant (HD) subphenotypes in sickle cell disease.

## Discussion

Our study achieved its primary goal of developing a methodical and feasible ML pipeline to analyze a sizable international SCD clinical trial dataset. By integrating disease pathophysiology with computational methods, we identified clinical subphenotypes and laboratory biomarkers, laying the groundwork for future predictive disease modeling (Figure 1). Using a combined MCA and K-means clustering approach, we uncovered hidden baseline patterns in clinical features of SCD and identified two distinct subphenotypes. The first, the hemolysis dominant (HD) subphenotype, is defined by the presence of hemolytic clinical features, including PHT, stroke, priapism, and ulcers—all of which clustered together in our model, suggesting a shared underlying pathophysiological process (Figure 2C). This subphenotype exhibited significantly lower Hb levels and elevated markers of hemolysis, such as LDH and indirect bilirubin (Figure 3, Table 1). In contrast, the vasoocclusive primary (VP) subphenotype was characterized by a higher annual rate of VOC (Figure 4) and absence of any hemolytic clinical features (Figure 2C). The VP subphenotype exhibited significant higher Hb, HbF and leukocyte counts (Figure 3, Table 1). These patterns were further validated using the independent clinical study, MSH. Our findings underscore the clinical and pathophysiological heterogeneity of SCD. Hemolysis plays a direct role in complications such as PHT, stroke, priapism, and ulcers through mechanisms involving vascular dysfunction, increased blood viscosity, and inflammation. In contrast, complications like pain crises, ACS, and AVN, are more directly linked to vascular occlusion and ischemic injury due to hemoglobin S polymerization and red cell sickling. Understanding these distinct pathophysiological pathways can inform clinical trial design and support the development of personalized treatment plans tailored to each subphenotype.

Kato et al. previously hypothesized two SCD subphenotypes based on pathophysiology and literature review: (1) the viscosity vasoocclusive subphenotype characterized by VOC, ACS, and AVN; (2) hemolytic endothelial dysfunction subphenotype marked by ulcers, PHT, priapism and stroke (15, 23, 29, 31). Our rule-based ML classification derived from clinical trial data supports the general structure of Kato’s hypothesis. However, while Kato proposed bidirectional overlap between the two subphenotypes (23), our findings reveal a more unidirectional pattern. In our model, hemolytic features (PHT, priapism, stroke, and ulcers) are exclusively clustered within the HD subphenotype (Figure 2C and 2E Supplementary Figure 5). In contrast, vaso-occlusive features of VOC, ACS and AVN appeared in both subphenotypes, but with small differences in distribution: 65.9%, 23.5%, 21% in the VP subphenotype versus 60.4%, 24.6%, 27.3% in the HD subphenotype, respectively (Supplementary Table 2, Supplementary Figure 5). Although the differences in VOC and AVN distribution were statistically significant, these differences were relatively small. These findings suggest that hemolytic features play a more decisive role than vaso-occlusive features in driving our ML-based subphenotype classification. Unlike Kato’s’ bidirectional model, our results suggest a unidirectional overlap—vaso-occlusive features appear in both subphenotypes, but hemolytic features are largely confined to HD. These discrepancies may stem from differences in data source, granularity, sample size, and methodology between the two studies, warranting further investigation.

Laboratory biomarker profiling in our rule-based subphenotypes model is aligned with the findings of Kato et al (23). In our primary analysis using the clinical trial database, the mean Hb was significantly higher in the VP subphenotype compared to the HD subphenotype (although only by 0.3 g/dL, p < 0.001) This trend was also observed in the secondary analysis using the MSH study data, with a difference of 0.45 g/dL (p = 0.002, Figure 3H, 5G). These findings align with the clinical intuitions, that higher Hb and HbF in the VP subphenotype could be the result of the body’s compensatory mechanisms to address ischemia and pain. Similarly, Kato et al. reported that higher Hb levels were associated with increased risk of vaso-occlusive complications (VOC, AVN, and ACS), whereas lower Hb levels are associated with hemolytic complications (ulcers, PHT, priapism and stroke) (23). Further validation of these predictive marker patterns in a larger SCD database is warranted.

Higher levels of HbF can interfere with HbS polymerization, serving as a protective factor against vaso-occlusion. In our study, the VP subphenotype demonstrated significant higher mean HbF values in both primary (clinical trial database) and secondary (MSH data) biomarker analyses (Figure 3H, 5G). However, the magnitudes of the difference in mean HbF between HD and VP subphenotypes varied: 0.8% in the clinical trial database vs. 1.5% in the MSH data. This discrepancy may be due to a substantial proportion of missing baseline HbF data in our clinical trial database – 43% in the VP subphenotype and 38.3% in the HD subphenotype (Supplementary Table 4). In contrast, the MSH data had no missing baseline laboratory values.

Another possible reason for the relatively higher mean HbF observed in the pooled clinical trial data compared to the MSH study may be due to the age distribution difference between the two datasets. The clinical trial database included 17.5% (n=506) patients younger than 10 years, and 37% (n=1,074) younger than 18 years (Supplementary Table 1), whereas MSH participants were adults aged 18-54 years (Supplementary Table 5). This age disparity could explain the HbF differences, as HbF concentrations in patients with SCD are influenced by age, sex and the geographical distribution of genetic modifiers (2, 39). In individuals with SCD, HbF switching is delayed to 5-10 years of age (40), whereas in the normal population HbF declines approximately from 85% at birth to <1% at one year of age (14).

In addition, all patients enrolled in the MSH study were hydroxyurea naïve; whereas, our clinical trial database included patients with varying baseline hydroxyurea exposure. Specifically, 1,270 patients (44.0%) were hydroxyurea naïve, 1,479 (51.2%) were receiving stable hydroxyurea treatment, and 138 (4.8%) had missing hydroxyurea exposure data (Supplementary Table 1). Therefore, the smaller HbF difference observed between the HD and VP subphenotypes in the clinical trial database compared to the MSH study may be partly explained by the fact that over half of the patients (51.2%) in our clinical trial database were receiving hydroxyurea treatment at baseline.

Another notable finding observed in both subphenotypes is that the mean HbF value in patients with the HbSS genotype is significantly lower than in those with an Sβ-thalassemia genotype (Supplementary Table 7). Interestingly, among patients with an Sβ-thalassemia genotype, the mean HbF trended higher in the HD subphenotype compared to the VP subphenotype. In contrast, for patients with the SS genotype, the mean HbF trended lower in the HD subphenotype than in the VP subphenotype (Supplemental Table 7).

Although these findings align with clinical observations in SCD and support our rule-based classification, we were unable to definitively characterize whether patients with a primarily vaso-occlusive presentation tend to have higher HbF than patients with the hemolytic presentation using our current subphenotype models. This limitation is partly due to the parametric comparison method employed, where large variability in HbF values affected our ability to detect a consistent difference between subphenotypes.

Moreover, the clinical trial data used in this study lacked comprehensive molecular genetic information for all subjects. The foundational observation of the protective role of higher HbF in young children with SCD, first described by Janet Watson (41, 42), has driven the field toward increasingly refined genetic characterization of HbF gene expression in SCD. Over several decades, this has evolved into extensive subject-level genetic data collection-encompassing haplotypes, single-nucleotide polymorphism (SNP) or genome-wide association studies (GWAS), including regulatory elements such as X-or other chromosomal transcription factors that are associated with SCD (14, 43–75). For instance, HbF values <10% driven by certain SNPs or GWAS loci have been associated with increased risk of cerebral vasculopathy in children with SCD (76). Without incorporating patient level molecular genetic data into our database, we were unable to adequately examine the relationship between HbF values and observed genotypic and phenotypic subgroups. Further, the ML subphenotypes could potentially be refined with additional hemolytic markers and subject level genetic data on deletional or nondeletional α-thalassemia (77–82). Further clinical trials incorporating this level of genetic detail could significantly enhance the precision of subphenotype classification in SCD.

Aligning with Kato’s hypothesis, findings from our study revealed that the VP subphenotype had a significantly higher mean HbF value and a significantly higher mean leukocyte count compared to the HD subphenotype (Figure 3 and Table 1). Further analyses revealed that HbF values were inversely correlated with absolute neutrophil and leukocyte counts in both VP and HD subphenotypes (Supplementary Figure 6). This finding is consistent with a report on increases in cumulative level of HbF silencing factors secreted by leukocytes that suppress gamma-globin gene expression (83). In addition, our analysis did not identify correlations between HbF value and the annual rate of VOC, indirect bilirubin or LDH in either subphenotype. An exception was noted in the VP subphenotype, where HbF was inversely correlated with reticulocyte count (Supplementary Figure 6). These results aligns with previous literature reports that increased HbF values are associated with a reduced frequency of VOC events, and fewer hemolysis related complications, such as an absence of leg ulcers in the Arabian Indian haplotype (53), but do not significantly influence hemolytic makers in other patients such as those with the severe HbS D-Punjab genotype (84).

The findings of older age of our HD subphenotype compared to the VP subphenotype is expected, (Figure 2D and Supplementary Figure 2), given that the HD subphenotype was defined by clinical conditions such as PHT, stroke, ulcers, and priapism, which are clinical conditions positively correlated with age. However, this age discrepancy introduces a potential confounding factor in our biomarker analysis, as many biomarkers are age dependent. To account for this, we conducted GLM analysis with the formula: biomarker ∼ subphenotype + age. This analysis confirmed that age is significantly associated with most biomarkers in our database. Nevertheless, subphenotypes remained significant for associations with Hb, LDH, and a non-significant, but consistent trend, with indirect bilirubin (Supplementary Table 6), suggesting the relevancy of that subphenotypes in biomarker differences overweighing the impact of age.

Although SCD is a rare disease, our study is distinguished by the large clinical trial database that we curated and assembled from 16 selected clinical trials. This resource enabled development of our ML pipeline and facilitated a more comprehensive exploration of clinical patterns in SCD, thereby enhancing the robustness and generalizability of our findings. However, when compared to common diseases, even a cohort of several thousands of patients does not constitute true ‘big data’. Further, missing baseline laboratory values (Figure 2E, Supplementary Table 4) presented challenges for applying more complex data mining techniques to our data.

Despite these limitations, the ML model developed in our analysis effectively tackled several common challenges encountered in rare disease datasets. Principal Component Analysis (PCA) (85) combined with K-means is among the most commonly used approaches for clustering. However, both PCA and K-means are sensitive to data scaling and missing values. To address these limitations in cluster modeling in our disease specific data, we made several modifications. First, we selected clinical features with the least missing data, allowing us to retain the largest possible patient cohort for analysis. Second, we replaced PCA with Multiple Correspondence Analysis (MCA), which is more suitable for categorical data. Beyond dimensionality reduction, MCA generated continuous numeric coordinates that captured the relationships among binary clinical features associated with SCD. This was particularly advantageous because K-means clustering requires numeric input, and MCA provided a robust and algorithmic means to transform categorical features accordingly. Using the clinical feature clusters generated by our modified ML pipeline, we assigned patients into their perspective subphenotypes. We then explored each biomarker independently, excluding patients with missing data for that specific variable. Rather than imputing missing values– particularly when insufficient information is available– we chose to preserve data integrity and minimized potential bias. Overall, our integration of ML techniques and statistical methods offers a novel pipeline for exploring clinical datasets with mixed data structure and limited sample sizes. This framework enhances the characterization of SCD subphenotypes and provides a valuable methodological path for research in rare diseases.

In addition to our subphenotypes described, we explored supervised learning approaches to integrate demographics, clinical features, biomarkers, and genetic data to predict SCD subphenotypes. However, due to the database size limitation and the high proportion of missing data, we were unable to impute missing values reliably, which precluded the effective use of Random Forest (86). As an alternative, we applied XGBoost (87), which achieved a high F1 score for the VP subphenotype. In contrast, performance for the HD subphenotype –representing less than 20% of the study population - was suboptimal, with an unsatisfactory maximal F1 score of approximately 0.3. We also attempted unsupervised visualization using UMAP (88). However, the binary nature of most variables, combined with the substantial missingness in biomarker data, limited our ability to identify meaningful or distinct patterns.

As discussed, our study has limitations. While the clinical trial database we assembled is relatively large for an orphan disease, the sample size remains modest for ML applications, which restricted the use of certain methods. Additionally, the substantial amount of missing data led to exclusion of 18.7% patients from the analysis due to incomplete clinical features (Figure 1). Among the 2,887 patients included in the analysis, missing laboratory data ranged from 9.2% to 44.5% across biomarkers (Supplementary Table 4). The precision of ML-based subphenotype classification was constrained by both limited sample size and the extent of missing data. Furthermore, the range of usable variables available for subphenotype classification in the clinical trial database was restricted to just 14 variables – comprising 7 clinical features and 7 laboratory biomarkers.

We applied our rule-based subphenotype classification to an independent clinical study, the MSH study, to evaluate biomarker differences between subphenotypes. The patterns of Hb, HbF, and indirect bilirubin levels observed in the MSH study were consistent with those observed in the clinical trial database. However, several limitations associated with the MSH data analysis should be noted. First, our subphenotype classification pipeline did not perform satisfactorily on MSH data, primarily due to an incomplete clinical feature set -only 5 out of 7 clinical features were available. In addition, LDH levels were not available, and the granularity of baseline VOC data was insufficient. Second, the small sample size (N = 299) further limited the robustness of our analysis. Third, after assigning subphenotypes to patients with incomplete clinical feature data, the VP to HD ratio in the MSH dataset was 1.25:1, compared to 4.22:1 in our clinical trial database. This shift suggests a relative overrepresentation of HD-classified subphenotypes in the MSH data. One possible explanation is that all participants in the MSH study were over 18 years old (Supplementary Table 5), making them more likely to develop age-associated complications such as stroke, leg ulcers, and priapism – features linked to the HD subphenotype. We did not observe significant differences in indirect bilirubin, leukocytes, and the annual rate of VOC between subphenotypes from MSH data, likely due to the limited sample size. Furthermore, the MSH study inclusion criteria required an annual rate of VOC ≥ 3, effectively excluding patients with milder disease. As a result, comparison between VP and HD subphenotypes in MSH data may be confounded by the inclusion of mixed or misclassified phenotypes. This sub-optimal classification likely contributed to the lack of statistically significant differences in certain biomarkers and annual rate of VOC between subphenotypes (Figure 4, Supplementary Figure 4).

Another potential limitation lies in our choice of classification pipeline. While MCA is a well-established and widely used method for analyzing binary variables, it is not the only option. More advanced ML algorithms, including deep learning approaches, may offer improved classification performance. However, as previously discussed, the limited size of our clinical trial database and the high proportion of missing data make it challenging to implement and validate such complex models. The finding of our study should be further validated using a larger database with less missing, more complete data. Further study should also focus on linking subphenotypes to clinical outcomes. This will be a crucial step toward developing predictive ML models that can enhance disease understanding, enhance prognosis, and ultimately enable personalized treatment strategies for patients with SCD. By integrating clinical characteristics, genetic information, and laboratory results, such a model could guide and support more targeted and effective interventions.

## Data Availability

Data are not available to public.

## Acknowledgement

This article reflects the views of the authors and should not be construed to represent views or policies of the FDA.

We wish to express our thanks to the following individuals and institutes:

1. Dr. Ann Farrell for her leadership, insightful guidance, and support for the project during her tenure.
2. Dr. Tanya Wroblewski for her leadership, insightful guidance, and support for the project and manuscript preparation.
3. Dr. Sergey Rakhilin, for providing specific materials to this project.
4. NHLBI for generously sharing their MSH database.

## Institutional review board statement

### Author contributions

Conceptualization, Study design and clinical implication: Q.R. and P.O.

Data curation: N.M., Q.R., R.M.Z., and W.X.

Methodology, Analytical design: M.W., Q.L., W.X., and Q.R.

Initial Analysis: M.W.

Formal analysis and Writing—original draft: W.X. and Q.R.

Contributing, writing—review & editing: All authors had full access to all the data related to this study and had final responsibility for the decision to submit for publication.

Supervision, Final editing, Resource, and Funding acquisition: Q.R., and P.O.

### Declarations

No competing or financial interest.

---

**Supplementary Figure 1.**
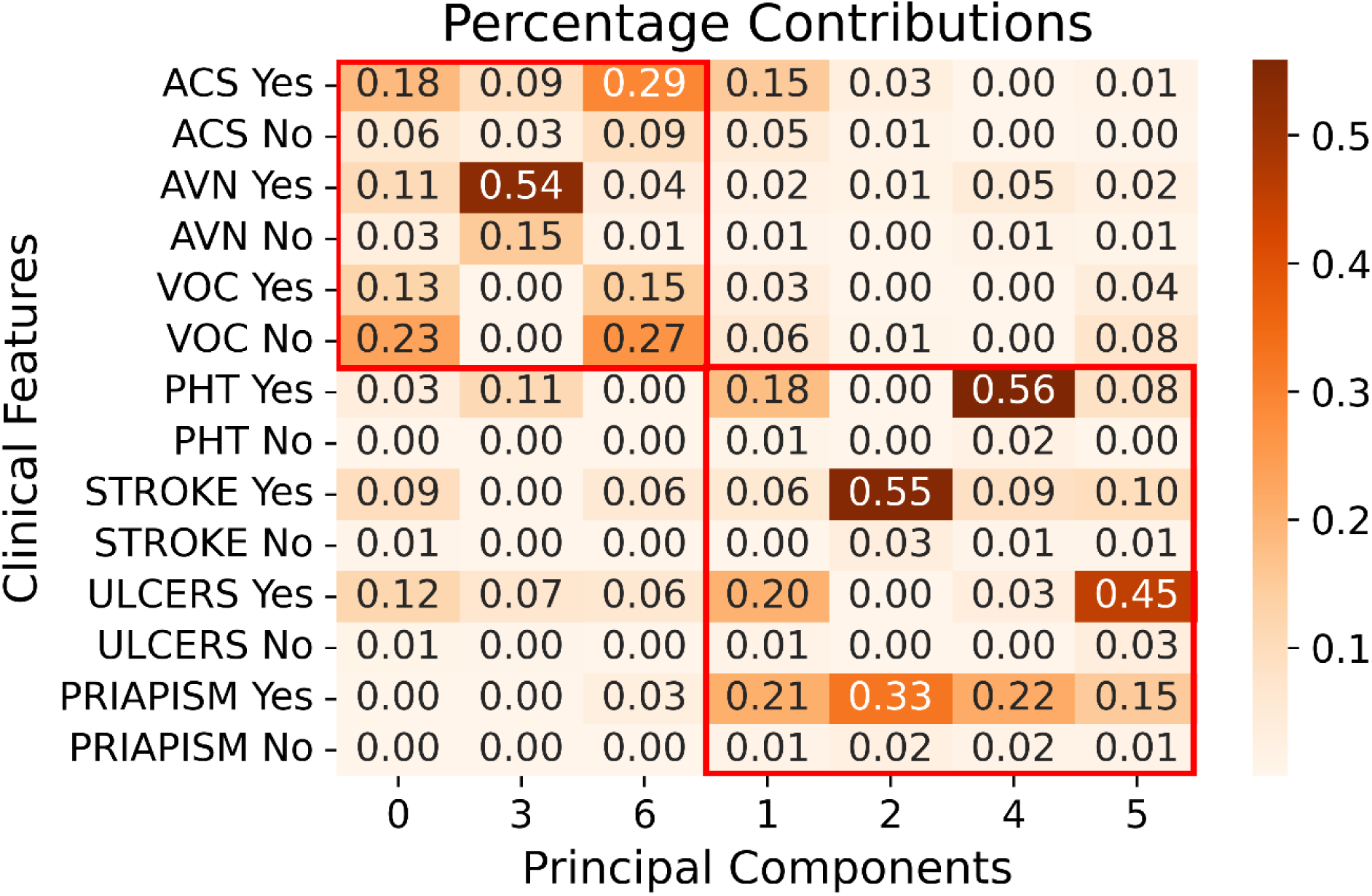
The percentage contribution of each category (“Yes” or “No”) within each clinical feature to each principal component.

**Supplementary Figure 2.**
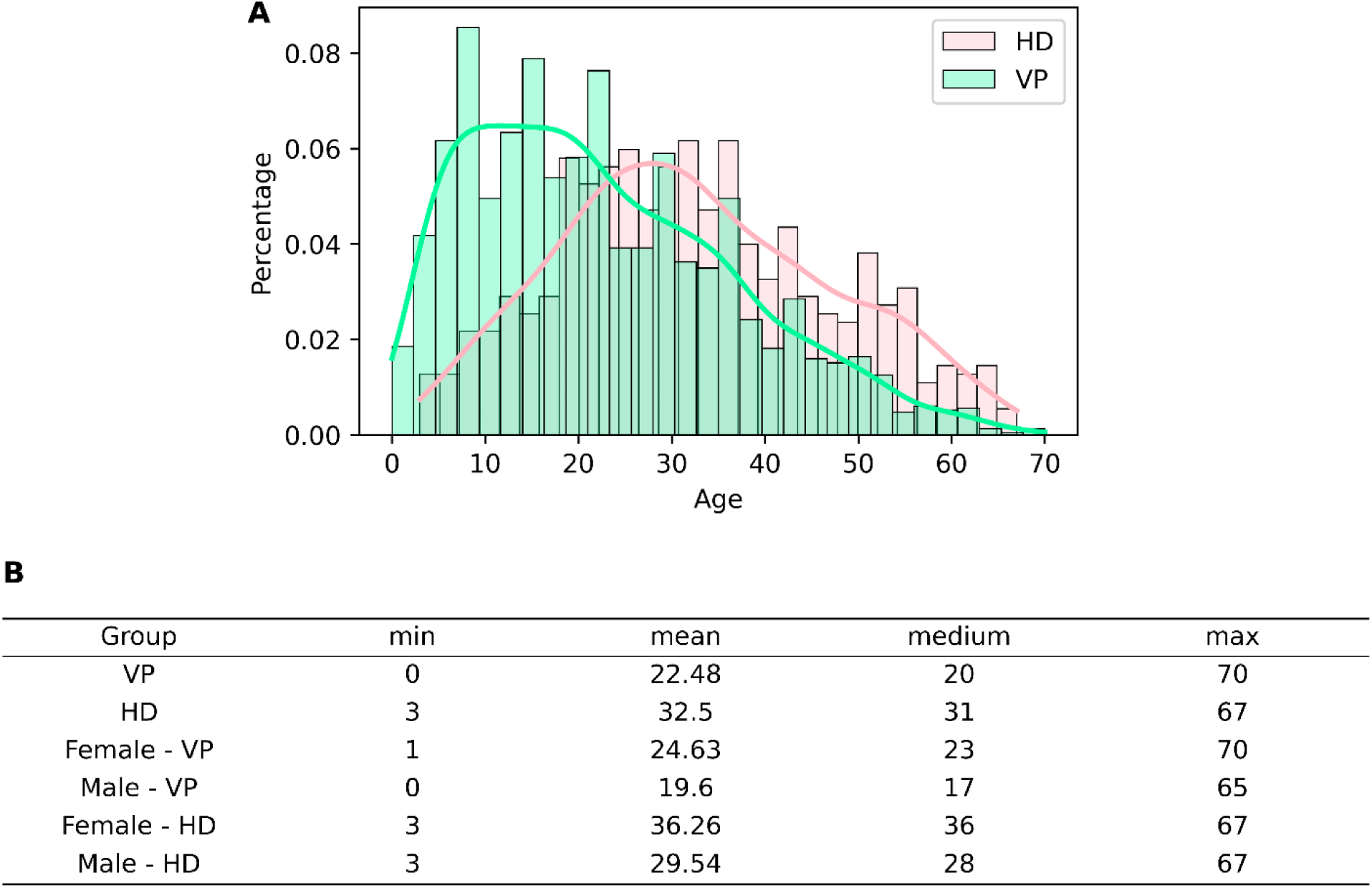
Age distributions across gender and subphenotype subsets. (A) Percentage histograms of age distributions for the two subphenotypes, with density curves overlaid. (B) The summary statistics table for each subset shown in Panels A and Figure 2D. VP: Vasoocclusive Primary subphenotype; HD: Hemolysis-Dominant subphenotype.

**Supplementary Figure 3.**
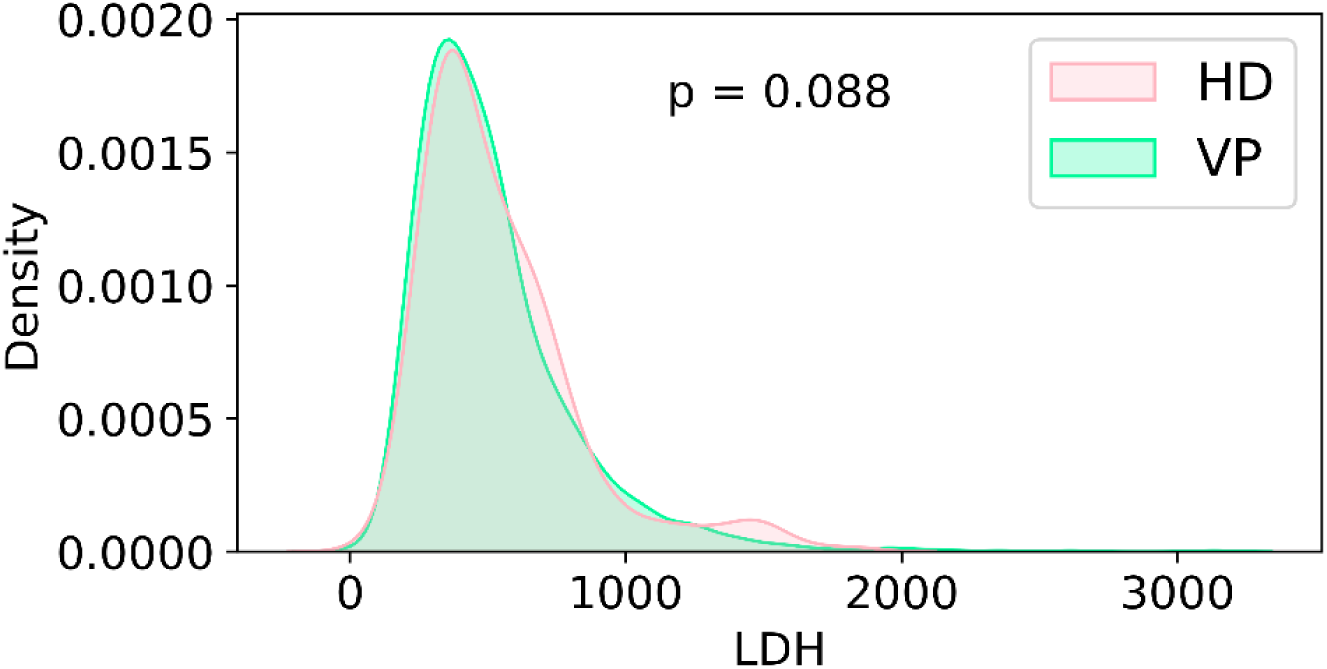
Density distribution of LDH by subphenotype (VP and HD) (U/L, N_total_=1,623, N_VP_=1,295, N_HD_=328). A bimodal distribution in HD is clearly visualized. Mann-Whitney U test p value is displayed. VP: Vasoocclusive Primary subphenotype; HD: Hemolysis-Dominant subphenotype.

**Supplementary Figure 4.**
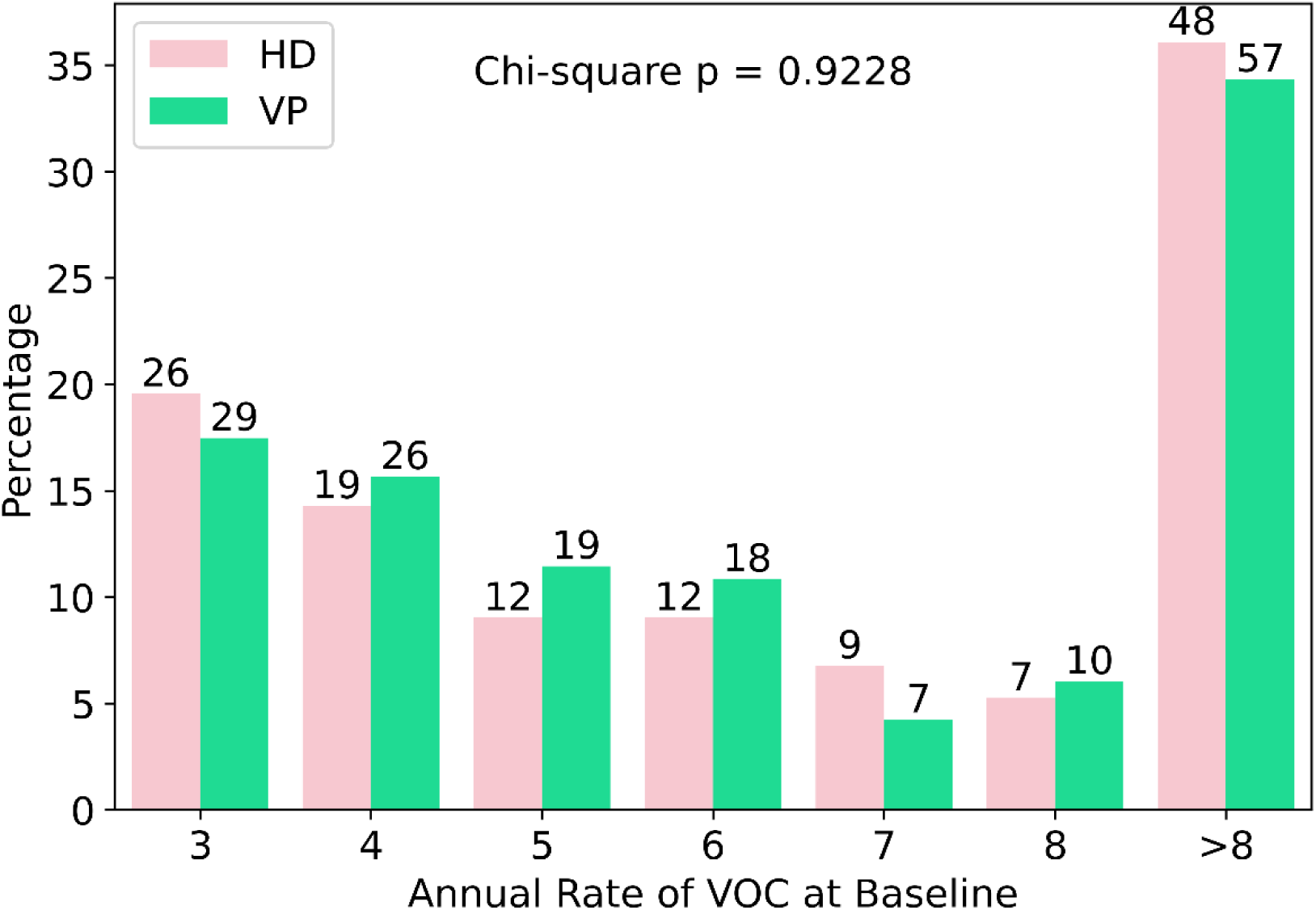
The figure shows the percentage of patients in each number of annual rate of VOC events at baseline, stratified by subphenotype of the MSH study. The patient counts in each group are labelled at the top of the bar. N_Total_ = 299, N_VP_=166, N_HD_=133. VP: Vasoocclusive Primary subphenotype; HD: Hemolysis-Dominant subphenotype.

**Supplementary Figure 5.**
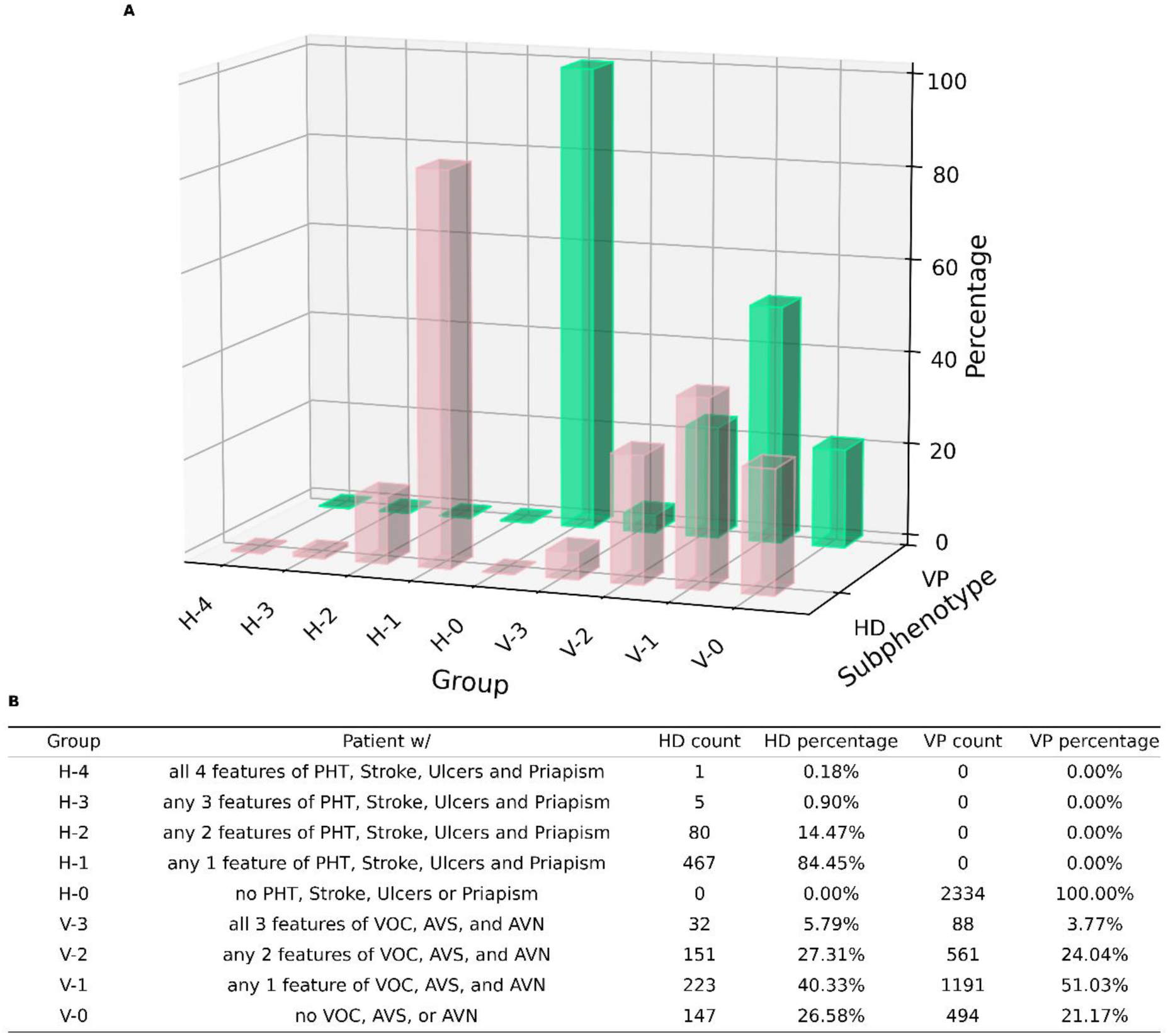
Clinical feature distribution across patients within subphenotypes. (A) A 3D bar plot illustrating the proportion of patients in each clinical feature combination group within the subphenotype. (B) A table detailing the group name, an explanation of the group’s clinical feature combination characteristics, and the patient count and percentage of groups within each subphenotype. VP: Vasoocclusive Primary subphenotype; HD: Hemolysis-Dominant subphenotype.

**Supplementary Figure 6.**
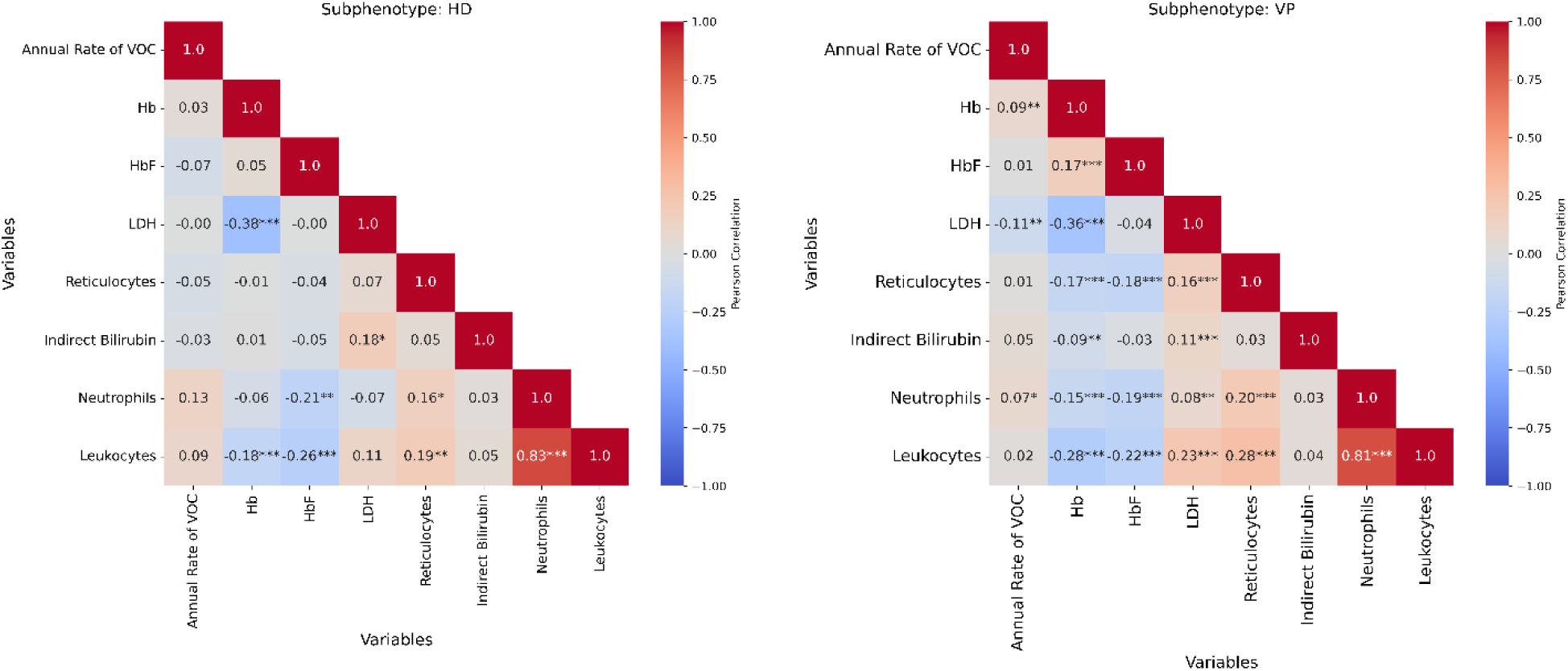
Correlation matrices of the clinical feature and laboratory biomarkers for HD and VP Subphenotypes. (A) and (B) display the Pearson correlation coefficients between the clinical feature, annual rate of VOC, and all available laboratory biomarkers for the HD and VP subphenotypes, respectively. The lower triangular heatmaps show the correlation matrix, with correlation coefficients colored and labeled in plot, and significant p-values indicated by asterisks (p < 0.001: ***, p < 0.01: **, p < 0.05: *). VP: Vasoocclusive Primary subphenotype; HD: Hemolysis-Dominant subphenotype.

**Supplementary Table 1.** Summary of SCD population from clinical trial database.

| **Population** | **Counts** | **Percentage** |
| --- | --- | --- |
| Number of Patients | 2887 | 100.00% |
| **Sex** |  |  |
| Female | 1576 | 54.59% |
| Male | 1300 | 45.03% |
| Unknown | 11 | 0.38% |
| **Race** |  |  |
| African American or Black | 725 | 25.11% |
| Other | 242 | 8.38% |
| Unknown | 1920 | 66.51% |
| **Age (year)** |  |  |
| 0-9 | 506 | 17.53% |
| 10-19 | 727 | 25.18% |
| 20-29 | 655 | 22.69% |
| 30-39 | 511 | 17.70% |
| 40-49 | 279 | 9.66% |
| 50-59 | 153 | 5.30% |
| 60+ | 42 | 1.45% |
| Unknown | 14 | 0.48% |
| Mean | 24.41 | 0.85% |
| Median | 22 | 0.76% |
| **Continent** |  |  |
| Europe | 2041 | 70.70% |
| North America | 412 | 14.27% |
| Africa | 102 | 3.53% |
| Asia | 68 | 2.36% |
| South America | 67 | 2.32% |
| Other | 59 | 2.04% |
| Unknown | 138 | 4.78% |
| **Pre-Hydroxyurea Treatment** |  |  |
| Yes | 1479 | 51.23% |
| No | 1270 | 43.99% |
| Unknown | 138 | 4.78% |

**Supplementary Table 2.** The Proportion of Subjects with Vasooclusive Features. This table presents the proportion of patients who have the listed clinical features (VOC, ACS, AVN) at baseline in the VP and HD subphenotypes. P-values from Chi-square tests are shown to assess the statistical significance of differences between subphenotypes. N_VP_ = 2,334, N_HD_ = 553. VP: Vasoocclusive Primary subphenotype; HD: Hemolysis-Dominant subphenotype.

| Feature Name | Proportion of Subjects with<br>Vasooclusive Features in Each<br>Subphenotype |  | p value |
| --- | --- | --- | --- |
|  | VP (%) | HD (%) |  |
| VOC | 65.94 | 60.4 | 0.0162 |
| ACS | 23.52 | 24.59 | 0.6335 |
| AVN | 20.95 | 27.31 | 0.0015 |

**Supplementary Table 3.** Age-specific value ranges used for the normalization of laboratory biomarkers. These ranges were based on the literature reported for both normal individual and patient’s with SCD.

|  | Age group | Value range |
| --- | --- | --- |
| Hb | 0-1 | 0-17 |
|  | 1-17 | 0-15 |
|  | >=18 | 0-18 |
| HbF | 0-1 | 0-20 |
|  | 1-10 | 0-15 |
|  | 10-17 | 0-10 |
|  | >=18 | 0-5 |
| LDH | 0-1 | 0-450 |
|  | 1-17 | 0-370 |
|  | >=18 | 0-225 |
| Indirect bilirubin | - | 0-0.8 |
| Reticulocytes | - | 0-100 |
| Neutrophils | 0-1 | 0-15 |
|  | 1-17 | 0-10 |
|  | >=18 | 0-7 |
| Leukocytes | 0-1 | 0-19 |
|  | >1 | 0-11 |

**Supplementary Table 4.** Summary of missing data. Patient counts and percentages of missing data counts for biomarkers in the VP and HD subphenotypes. VP: Vasoocclusive Primary subphenotype; HD: Hemolysis-Dominant subphenotype.

| Biomarker/Clinical Feature | Missing data in VP | Missing data in HD |
| --- | --- | --- |
| Hb | 250/10.71% | 51/9.22% |
| HbF | 1004/43.02% | 212/38.34% |
| Indirect Bilirubin | 793/33.98% | 153/27.67% |
| LDH | 1039/44.52% | 225/40.69% |
| Reticulocytes | 806/34.53% | 167/30.20% |
| Neutrophils | 433/18.55% | 89/16.09% |
| Leukocytes | 284/12.17% | 58/10.49% |
| Annual Rate of VOC | 855/36.63% | 217/39.24% |

**Supplementary Table 5.**
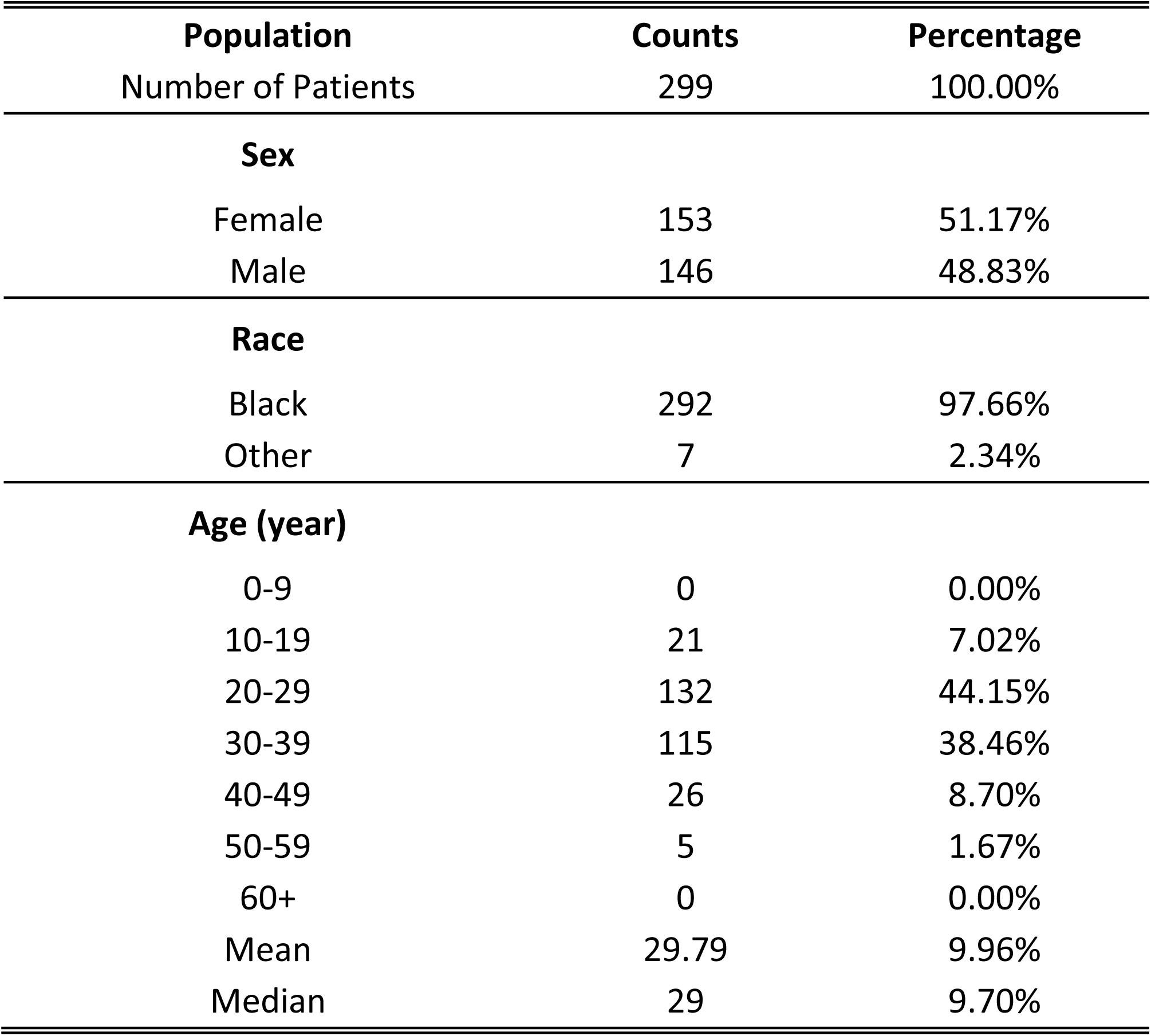
Summary of SCD population from MSH study.

| **Population** | **Counts** | **Percentage** |
| --- | --- | --- |
| Number of Patients | 299 | 100.00% |
| **Sex** |  |  |
| Female | 153 | 51.17% |
| Male | 146 | 48.83% |
| **Race** |  |  |
| Black | 292 | 97.66% |
| Other | 7 | 2.34% |
| **Age (year)** |  |  |
| 0-9 | 0 | 0.00% |
| 10-19 | 21 | 7.02% |
| 20-29 | 132 | 44.15% |
| 30-39 | 115 | 38.46% |
| 40-49 | 26 | 8.70% |
| 50-59 | 5 | 1.67% |
| 60+ | 0 | 0.00% |
| Mean | 29.79 | 9.96% |
| Median | 29 | 9.70% |

**Supplementary Table 6.** Coefficients and p-values from Generalized Linear Models (GLM) assessing the association between biomarkers and subphenotype and age. The table presents the estimated coefficients for the effect of subphenotype and age on each biomarker, along with the corresponding p-values indicating statistical significance. A positive coefficient for subphenotype indicates higher biomarker levels in VP patients compared to HD patients, adjusted for age. VP: Vasoocclusive Primary subphenotype; HD: Hemolysis-Dominant subphenotype.

| Biomarker | Subphenotype |  | Age |  |
| --- | --- | --- | --- | --- |
|  | Coefficient | p value | Coefficient | p value |
| Hb | 0.43 | < 0.001 | 0.01 | < 0.001 |
| HbF | 1.1 | 0.059 | 0.03 | 0.058 |
| LDH | -60.11 | 0.001 | -3.78 | < 0.001 |
| Indirect Bilirubin | -0.83 | 0.083 | -0.04 | 0.004 |
| Reticulocytes | -29.29 | 0.012 | -2.32 | < 0.001 |
| Leukocytes | -0.42 | 0.032 | -0.08 | < 0.001 |
| Neutrophils | -0.04 | 0.804 | -0.02 | < 0.001 |

**Supplementary Table 7.** Multiple Comparison of Means using Tukey HSD for the comparison of HbF between different genotypes (HbSBeta and HbSS) and subphenotypes (HD and VP) combinations. The p-values represent the adjusted significance levels (FWER = 0.05) for pairwise comparisons, with meandiff indicating the difference in means between the groups (Higher mean group is marked in bold, meandiff = mean(group2) - mean(group1)). A p-value less than 0.05 suggests a statistically significant difference. N_HD HbSBeta_= 33, N_HD HbSS_= 281, N_VP HbSBeta_= 230, N_VP HbSS_= 934. VP: Vasoocclusive Primary subphenotype; HD: Hemolysis-Dominant subphenotype.

| Multiple Comparison of Means - Tukey HSD, FWER=0.05 |  |  |  |
| --- | --- | --- | --- |
| Group 1 | Group 2 | p-adj | meandiff |
| **HD HbSBeta** | HD HbSS | **0.0168** | -5.0255 |
| **VP HbSBeta** | VP HbSS | **<0.001** | -3.1683 |
| **HD HbSBeta** | VP HbSBeta | 0.8884 | -1.2428 |
| HD HbSS | **VP HbSS** | 0.7629 | 0.6144 |

